# Development and evaluation of a novel educational program for providers on the use of polygenic risk scores

**DOI:** 10.1101/2023.02.16.23286057

**Authors:** T. Yanes, C. Wallingford, MA. Young, A.M. McInerney-Leo, A. Willis, L. McKnight, B. Terrill, S. McInerny, L. Forrest, L. Cicciarelli, R. Williams, H. Keane, PA James

**Affiliations:** Frazer Institute, The University of Queensland, Dermatology Research Centre, Brisbane QLD 4102; Clinical Translational and Engagement Platform, Garvan Institute of Medical Research, Darlinghurst, NSW, Australia; School of Clinical Medicine, UNSW Medicine & Health, St Vincent’s Healthcare Clinical Campus, Faculty of Medicine and Health, UNSW Sydney, NSW, Australia; Parkville Familial Cancer Centre, Peter MacCallum Cancer Centre and the Royal Melbourne Hospital, Melbourne VIC 3000, Australia; School of Clinical Medicine, UNSW Medicine & Health, UNSW Sydney, Kensington, NSW, Australia; Hereditary Cancer Centre, Prince of Wales Hospital, Randwick, 2031, Australia; Sir Peter MacCallum Department of Oncology, University of Melbourne, Melbourne VIC 3052, Australia

**Author notes:** Corresponding authors: Tatiane Yanes. Authors contributed equally (T. Yanes, C. Wallingford).

**Keywords:** breast cancer, ovarian cancer, education, polygenic risk scores

## Abstract

**Background:** Polygenic risk scores (PRS) for breast and ovarian cancer risk are increasingly available to the public through clinical research and commercial genetic testing companies. Healthcare providers frequently report limited knowledge and confidence using PRS, representing a significant barrier to evaluation and uptake of this technology. We aimed to develop and evaluate the impact of a novel online educational program on genetic healthcare providers (GHP) attitudes, confidence and knowledge using PRS for breast and ovarian cancer risk.

**Methods:** The educational program was informed by adult learning theory and the Kolb experiential learning model. The program was comprised of two phases: i) an online module covering the theoretical aspects of PRS and ii) a facilitated virtual workshop with pre-recorded role plays and case discussions. A pre-and post-education survey was administered to evaluate the impact of the educational program on GHP attitudes, confidence, knowledge, and preparedness for using PRS. Eligible participants were GHP working in one of 12 familial cancer in Australia registered to recruit patients for a breast and ovarian cancer PRS clinical trial and completed the education program.

**Results:** 124 GHP completed the PRS education, of whom 80 (64%) and 67 (41%) completed the pre- and post-evaluation survey, respectively. Pre-education, GHP reported limited experience, confidence and preparedness using PRS. GHP frequently recognized potential benefits to PRS, most commonly that this information could improve access to tailored screening (rated as beneficial/very beneficial by 92% of GHP pre-education). Completion of the education program was associated with significantly improved attitudes (p=<0.001), confidence (p=<0.001), knowledge of (p=<0.001) and preparedness (p=<0.001) using PRS. Most GHP indicated the education program entirely met their learning needs (73%) and felt the content was entirely relevant to their clinical practice (88%). GHP identified further PRS implementation issues including limited funding models, diversity issues, need for clinical guidelines and ongoing updates given the rapid pace of PRS research.

**Conclusions:** Delivery of a novel education program can improve GHP attitudes, confidence, knowledge, and preparedness using PRS. Careful consideration of healthcare providers’ learning needs is required to support PRS research and clinical translation.

## Introduction

Assessing breast cancer risk based on known risk factors, such as family history and age, is well established in clinical practice and the basis for most population screening programs (1). Separately, at the high end of familial risk, genetic testing is also well established for rare pathogenic/likely pathogenic variants (PV/LPV) in a panel of breast cancer genes (2). The best known are the *BRCA1* and *BRCA2* (*BRCA1/2*) genes, which convey a cumulative lifetime breast cancer risk of 50-70%, along with significant risks for ovarian cancer and other malignancies (3). For women who carry a PV/LPV in *BRCA1/2*, high quality evidence exists for the effectiveness of risk management options including breast screening, risk reducing medication, prophylactic mastectomy and bilateral salpingo-oophorectomy (4). Moderate penetrance breast cancer risk genes have also been identified including *ATM* and *CHEK2*, and genetic testing for high and moderate penetrant genes is now considered a standard part of the assessment of breast cancer care (5).

A complimentary advance in the understanding of breast cancer risk has been the identification of hundreds of common, low risk single nucleotide polymorphisms (SNPs) (5, 6). Although the effect of each SNP is very small, they can be combined into a single measure, known as a polygenic risk score (PRS) that has been found to effectively describe the distribution of breast cancer risk in the general population (7, 8) and in high-risk families (9-11). In contrast to rare variants, a PRS can be applied to all women and shows a normal distribution in the population (11). For any individual a PRS can be considered as the background risk, onto which all further risk factors are mapped (12).

Polygenic risk scores have been shown to be an independent risk factor to monogenic risk, and modify risk associated with high-and moderate-risk penetrance breast and ovarian cancer risk genes (13-17). In one study, the lifetime breast cancer risk for women with a *CHEK2* PV/LPV ranged from 14.3% to 32.6% for those with a PRS in the lowest and top quartile or risk distribution, respectively (14). More recently, the CanRisk tool (12, 18) has been validated to provide individuals with a personalized risk for breast and ovarian cancer based on established disease risk factors that includes: genetic information (i.e. monogenic and PRS), personal and family history of cancer, and clinical and lifestyle factors (e.g. reproductive history, breast density, alcohol consumption) (12). Despite the development of comprehensive risk prediction tools, the clinical, psychological and economic impact of providing individuals with their personalized breast and ovarian cancer risk remains unknown. It has been argued that providing personalized risk for women with PV/LPV in high penetrance genes is unlikely to alter risk management, as those individuals will remain at high risk regardless of the PRS and presence/absence of other risk factors. Conversely, providing this information may inform decisions-making regarding timing of risk-reducing surgeries.

Genetics healthcare providers are well equipped to oversee, interpret, and communicate monogenic risk information. However, PRS and personalized risk assessments represent a novel deviation from standard clinical genetic practice (19). For example, implementation of this information will require a shift towards a more personalized approach to healthcare, greater consideration of the multifactorial nature of disease risk, limitations of testing, and potentially re-assessment of risk over time (20). Genetic healthcare providers have reported limited knowledge and confidence regarding PRS and personalized risk assessment, and no formal training is available on how to use and interpret this information in clinical practice (21-23). The lack of knowledge among providers is a significant barrier to the successful evaluation and implementation of PRS and personalized risk assessment (11, 24).

### The PRiMo trial

The “Using Polygenic Risk Modification to Improve Breast Cancer Prevention” (PRiMo) is an Australian randomized clinical trial that aims to evaluate the health, economic and social impact of providing personalized breast and ovarian cancer risk information to women (25). Participants for the PRiMo trial are women referred to one of the 12 participating clinical genetic service in Australia for predictive testing of a breast and/or ovarian cancer risk gene (Supplementary Material 1). Women randomized to the intervention arm receive personalized breast and ovarian cancer risk information (generated from CanRisk; comprised of monogenic, polygenic, family, and lifestyle risk). The control group receives standard care, that includes only monogenic test results (25). It is anticipated that the evidence obtained from PRiMo will be used to inform the widespread implementation of personalized risk assessment in clinical practice.

The successful evaluation and implementation of personalized risk assessments is dependent on healthcare providers’ knowledge and confidence with this information (11, 24). Thus, the first phase of PRiMo included the development of a novel educational program designed to ensure that providers are confident with the theoretical and practical aspects of PRS for breast and ovarian cancer risk assessments, and that the PRiMO intervention is consistently delivered by multiple providers across study sites. The present study aimed to develop and evaluate the impact of a novel educational program on genetic healthcare providers’ attitudes, confidence, knowledge, and preparedness providing PRS and personalized risk assessment for breast and ovarian cancer.

## Methods

### Training development

Development of the education program was informed by adult learning theory and the Kolb Experiential learning model (26, 27). The education program was comprised of two phases: a website covering the theoretical aspects of PRS and personalized risk, and a virtual workshop focusing on risk communication (Supplementary Material 2). The website had five modules covering: i) PRS for Breast and Ovarian Cancer Risk, ii) Personalized Breast and Ovarian Cancer Risk, iii) Breast and Ovarian Cancer Risk Factors, iv) the PRiMo Trial, and v) Communicating Personalized Risk Information. The website was designed to facilitate reflection and development of abstract knowledge. As such, each module included: a video presentation, supplementary written information, recommended readings, true/false questions for self-knowledge reflection, and case studies with questions to facilitate self-reflection (https://learn.garvan.org.au/primo-clinician-training/). For participants who incorrectly answered a question, information on the correct response and recommended reading was provided.

Two weeks after receiving access to the website, providers participated in a 1.5-hour virtual workshop moderated by authors TY and PAJ (delivered via Zoom). The workshop included four pre-recorded roleplays designed to facilitate reflection and discussion regarding practical aspects of PRS and personalized risk information. Each case covered a different issue related to personalized risk: i) strategies for communicating complex risk information (i.e. monogenic, polygenic and other risk factors), ii) impact of personalized risk on cancer risk management, iii) communicating personalized risk in the context of familial testing, and iv) addressing diversity issues in genomics, PRS and healthcare.

Providers could self-select which online modules to complete prior to attending the virtual workshop based on their own knowledge gaps. However, participation in the subsequent virtual workshop was mandatory for all genetic healthcare providers recruiting for PRiMo. The educational program was pilot tested with 24 providers over the course of three workshops prior to widespread delivery. Based on participants’ feedback during the pilot stage, the virtual workshop was modified, namely re-recording one of the roleplays and adjusting the wording of reflective questions.

### Training delivery and evaluation

Delivery of the education program and data collection occurred between August 2021 and October 2022. Eligible genetic healthcare providers included clinical geneticists, genetic counsellors and medical oncologists who were working in one of the 12 familial cancer clinics in Australia registered to recruit patients for PRiMo. Eligible providers were identified by the local site investigator and their email address given to the PRiMo study coordinator (author SM).

An anonymous pre-post survey was developed to evaluate the effects of the education program on genetic healthcare providers’ general attitudes, perceived benefits and concerns, knowledge, confidence, and preparedness using PRS and personalized risk assessments. All eligible providers were sent an email with a link to the education website, information about the virtual workshop and an invitation to complete the pre-education survey (Supplementary Material 2). Information about the pre-education survey was also included in the homepage of the website. Immediately after the online workshop, a link to the post-education survey was provided to all attendees, with a reminder sent approximately 2-weeks post-training by the PRiMo coordinator (author SM). Providers were informed that they were eligible to complete the post-education survey regardless of whether they had completed the pre-survey.

The study information and consent form were available at the start of both surveys. Participation in the study and completion of the anonymous surveys was voluntary, and consent to participate was implied by completion of the survey. Both surveys were available through the University of Queensland Qualtrics Platform. At the start of each survey, participants were asked to put the last four digits of their mobile number and this information was used to match pre-and post-survey data. In addition to the survey data, de-identified data on website engagement was collected, including number of enrolled users, view count for video presentations, summary of time spent reviewing website content, and number of quizzes attempts.

### Survey development

Survey development was informed by the Kirkpatrick model, which comprises four criteria: reaction, learning, behavior and results (28). The first two criteria of the model were evaluated in this study. To evaluate learning and reaction, custom survey items were developed to assess the following outcomes: changes in general attitudes towards (22), perceived benefits and concerns, confidence, knowledge and preparedness related to the use of PRS and personalized risk in clinical practice (Supplementary Table 3). Total scores for attitudes, benefits, concerns, and confidence were calculated based on the sum of each item, with higher scores indicating more positive attitudes, and greater perceived benefits, concerns, and confidence. For the general attitudes, three negatively worded items were reversed coded (*this information will increase health disparities across ancestries, this information should not be provided to patients until there is sufficient data for people of all ancestries, and the thought of incorporating this information in my clinical practice scares me*), and a total score calculated. Knowledge was calculated based on the frequency of correct responses. Reaction was further captured in the post-education survey with participants providing feedback on their engagement with various aspects of the education program and usefulness of the information provided (29).

### Statistical analysis

Descriptive statistics, including frequencies and measures of central tendency, were used to describe the sample. Mean scores for general attitudes, benefits, concerns, confidence, and objective knowledge were calculated. Dependent sample t-test were conducted to evaluate changes pre-and post-education. Data from open-ended questions were analyzed using qualitative descriptive analysis.

## Results

### Engagement with education program

Over the course of the main study (i.e., excluding the pilot phase), the educational program was delivered to 124 providers across seven virtual workshops. Of the 124 eligible participants, 117 (94%) accessed the education website prior to the workshop, of whom 76 (64%) attempted all quizzes at least once, and 25 (9%) attempted a quiz more than once. The reflective case studies were submitted by 59 (50%) users. The median time spent on the website was 2 hours and 20 minutes (range: 11 minutes to 5.5 hours). Data on engagement with pre-recorded website presentations was obtained. The most frequently watched presentations were Module 1 “*Understanding PRS for Breast and Ovarian Cancer Risk*” (n=174 views), Module 2: *Understanding Personalized Breast and Ovarian Cancer Risk*” (n=163 views) and Module 5: “*Communicating Personalized risk*” (n=154 views). Of note, view counts included pilot study participants’ utilization.

### Participant demographic and experience with PRS

Of the 124 eligible providers, 80 (65%) completed the pre-education survey, and 67 (54%) the post-survey. Among participants, 52 (41%) completed both pre-and post-survey, and responses could be matched. Most participants were genetic counsellors (n=60, 73%), females (n=70, 85%), aged between 31-40 years old (n=31, 38%) and had between 0-5 years’ experience in clinical practice (n=41, 50%) (Table 1). Participants had little experience using PRS (n=44, 86% reporting no prior experience), and heard almost nothing (n=4, 5%) or some information on PRS (n=48, 60%). When asked about prior experience using personalized breast and ovarian cancer risk prediction, most (n=71, 89%) indicated having used the CanRisk tool to estimate risk cancer risk and eligibility for publicly funded genetic testing (i.e >10% chance of having PV/LPV (30)).

**Table 1:** Demographic characteristics of participant cohort and experience using PRS pre-education.

| **Item** | **N (%)** |
| --- | --- |
| *Age (n=82)* |  |
| 18-30 | 22 (27) |
| 31-40 | 31 (38) |
| 41-50 | 16 (20) |
| 51-60 | 11 (13) |
| 61 and above | 2 (2) |
| *Gender (n=82)* |  |
| Female | 70 (85) |
| Male | 12 (15) |
| *Profession (n=82)* |  |
| Genetic counsellor | 60 (73) |
| Clinical Genetics | 17 (21) |
| Oncologist | 5 (6) |
| *Years of practice (n=82)* |  |
| 0-5 | 41 (50) |
| 6-10 | 12 (15) |
| 11-15 | 12 (15) |
| 15-20 | 9 (11) |
| More than 20 years | 8 (10) |
| *How much have you heard about PRS (n=80)* |  |
| Almost nothing | 4 (5) |
| Some information | 48 (60) |
| A fair amount of information | 24 (30) |
| A lot of information | 4 (5) |
| *Have you every returned PGS? (n=80)* |  |
| No | 44 (86) |
| Yes, in research capacity | 5 (10) |
| Yes, in clinical capacity | 1 (2) |
| Yes, in research and clinical capacity | 1 (2) |
| *Experience using CanRisk (n=80)* |  |
| Yes, to estimate cancer risk | 4 (5) |
| Yes, to evaluate eligibility for publicly funded genetic testing | 1 (1) |
| Yes, to estimate cancer risk and evaluate eligibility for publicly funded genetic testing | 71 (89) |
| No, I do not use CanRisk | 4 (5) |

### Attitudes Towards PRS and Personalized Risk

Pre-education, participants were most likely to rate the following statements regarding PRS as strongly agree/somewhat agree: “*patients will appreciate receiving this information*” (n=67, 84%), “*this information should be a routine part of clinical care for familial breast and ovarian cancer*” (n=52, 65%) and “*this information is an accurate way to estimate cancer risk*” (n=50, 63%) (Figure 1). In relation to concerns about data diversity and health disparities, pre-education, a quarter of participants reported that “*PRS and personalized risk should not be provided until there is sufficient data for people of all ancestries*” (n=20, 25%). In comparison, post-education, only 2 (3%) participants strongly agree/somewhat agreed with this statement. Providers reported higher mean scores for general attitudes towards PRS and personalized risk after completing the education program (M=21.2, SD 1.5) compared to pre-education (M=12.5, SD=3.0) (t (49) = −23.3, p=<0.001).

**Figure 1:**
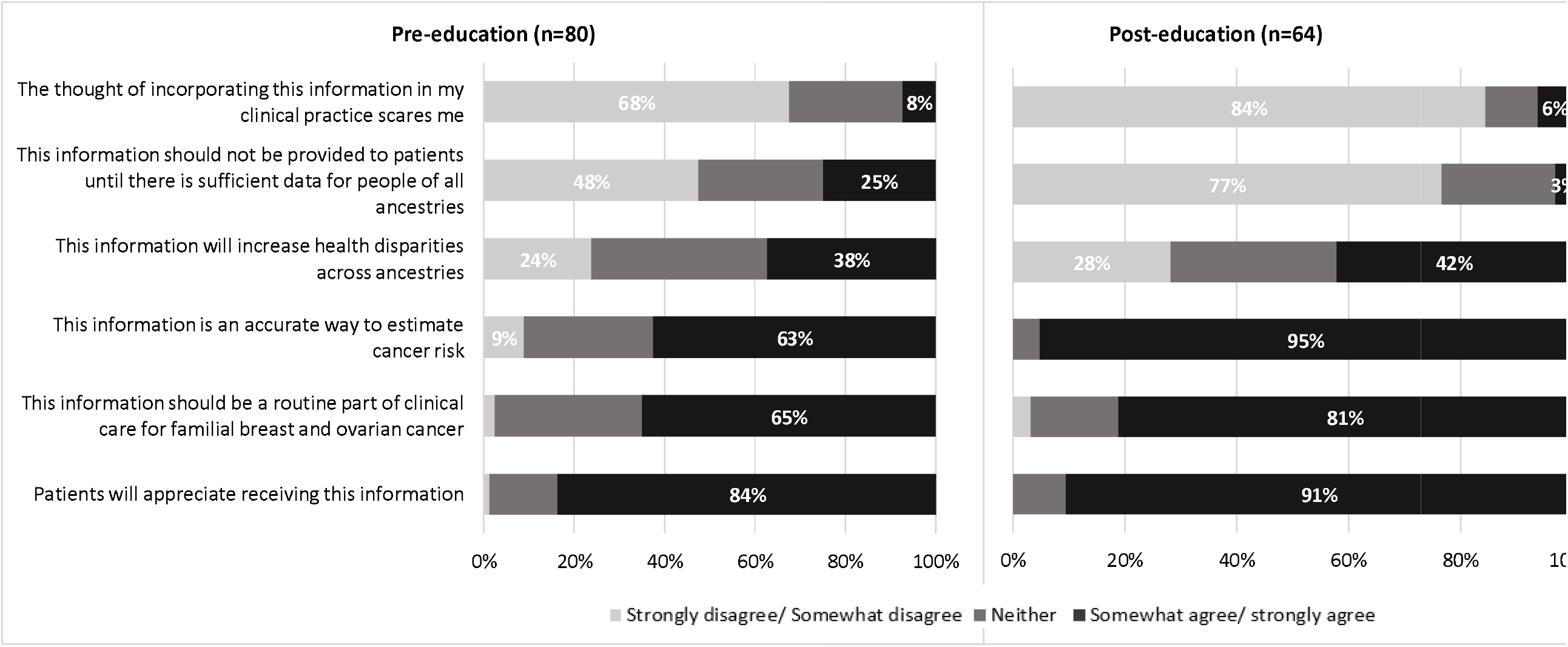
Changes in general attitudes towards breast and ovarian cancer PRS and personalised risk pre-and (n=80) post-education (n=64)

Pre-education, participants frequently identified potential benefits to providing PRS and personalized risk, with all items provided rated as beneficial/very beneficial by more than 60% of all participants (Figure 2). Pre-education, participants felt this information would improve access to tailored screening, provide more accurate information to make decisions about risk-reducing strategies, and provide greater certainty about level of cancer risk, rated as beneficial/very beneficial by 92%, 91% and 90% of participants, respectively. In relation to perceived concerns, participants most frequently identified the impact of ancestry on the interpretation of PRS, lack of national guidelines for PRS, and potential impact of yet to be identified SNPs on PRS, rated as concerns by 63%, 55% and 28% of participants, respectively (Figure 2). Overall, there was no change in total mean scores for perceived benefits pre- (M 2.6, SD 0.4) and post-education (M 3.0, SD 0.43) (t (46) = −1.1, p=0.26). Similarly, there was no change in total mean scores for reported concerns pre- (M 3.0, SD 0.4) and post-education (M 3.2, SD 0.47) (t (46)= 1.1, p=0.26).

**Figure 2:**
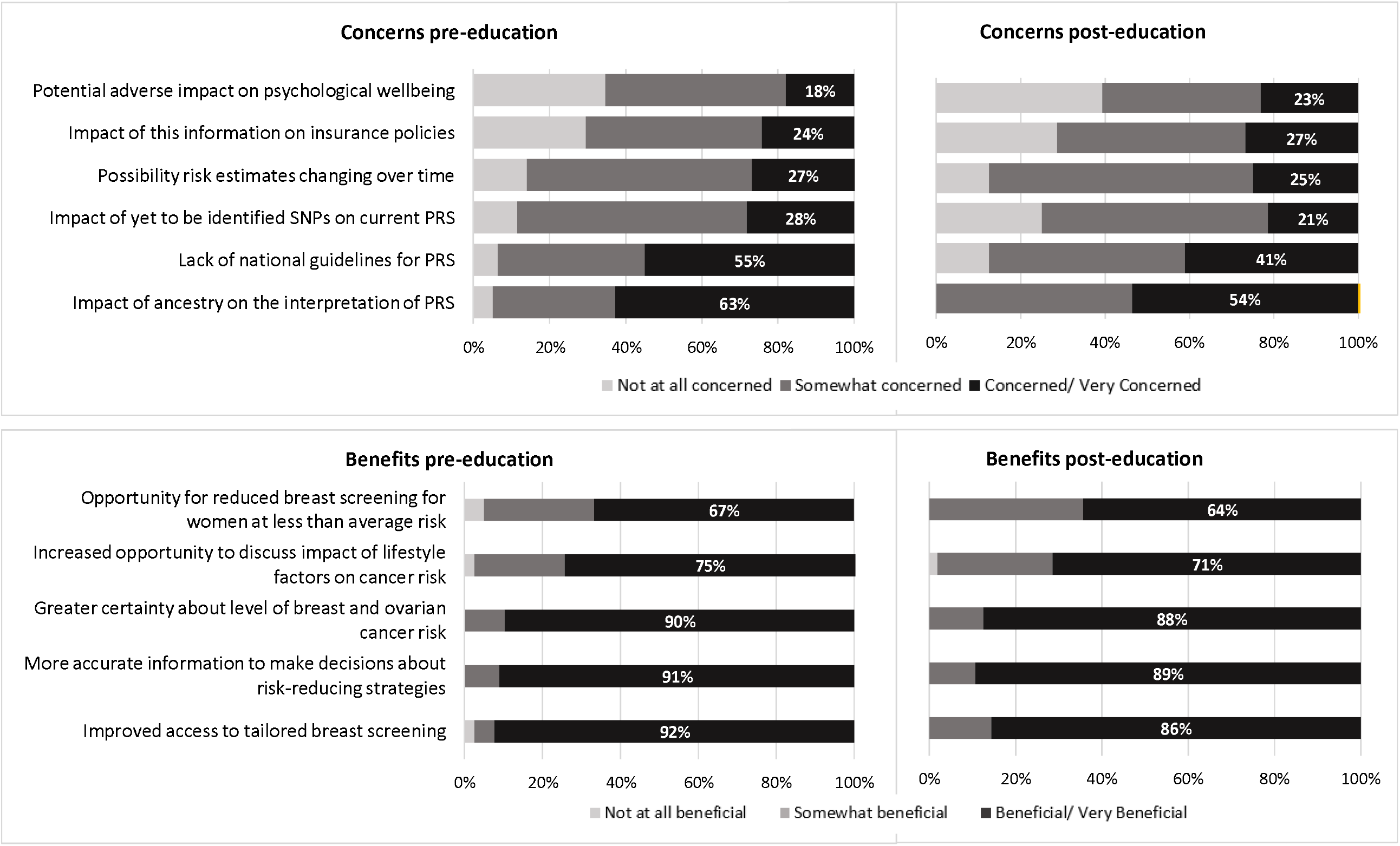
Changes in reported concerns and benefits of PGS and personalised risk pre-(n=78) and post-education (n=56)

Review of open text boxes identified several additional concerns described by providers, including patient acceptance of reduced screening within the public health system, impact of personalized risk within a familial setting, and capabilities of non-genetic healthcare providers to interpret and communicate PRS (Table 2). Participants also described implementation issues including a need to develop standardized guidelines that aim to mitigate further healthcare disparities, genetic health services not being able to meet additional demands for PRS and personalized risk implementation (such as a need for re-assessment over time), and limited funding model within the public health system to support PRS testing. Lastly, providers recognized a need for ongoing updates to any clinical guidelines given the rapid pace of PRS research.

**Table 2:**
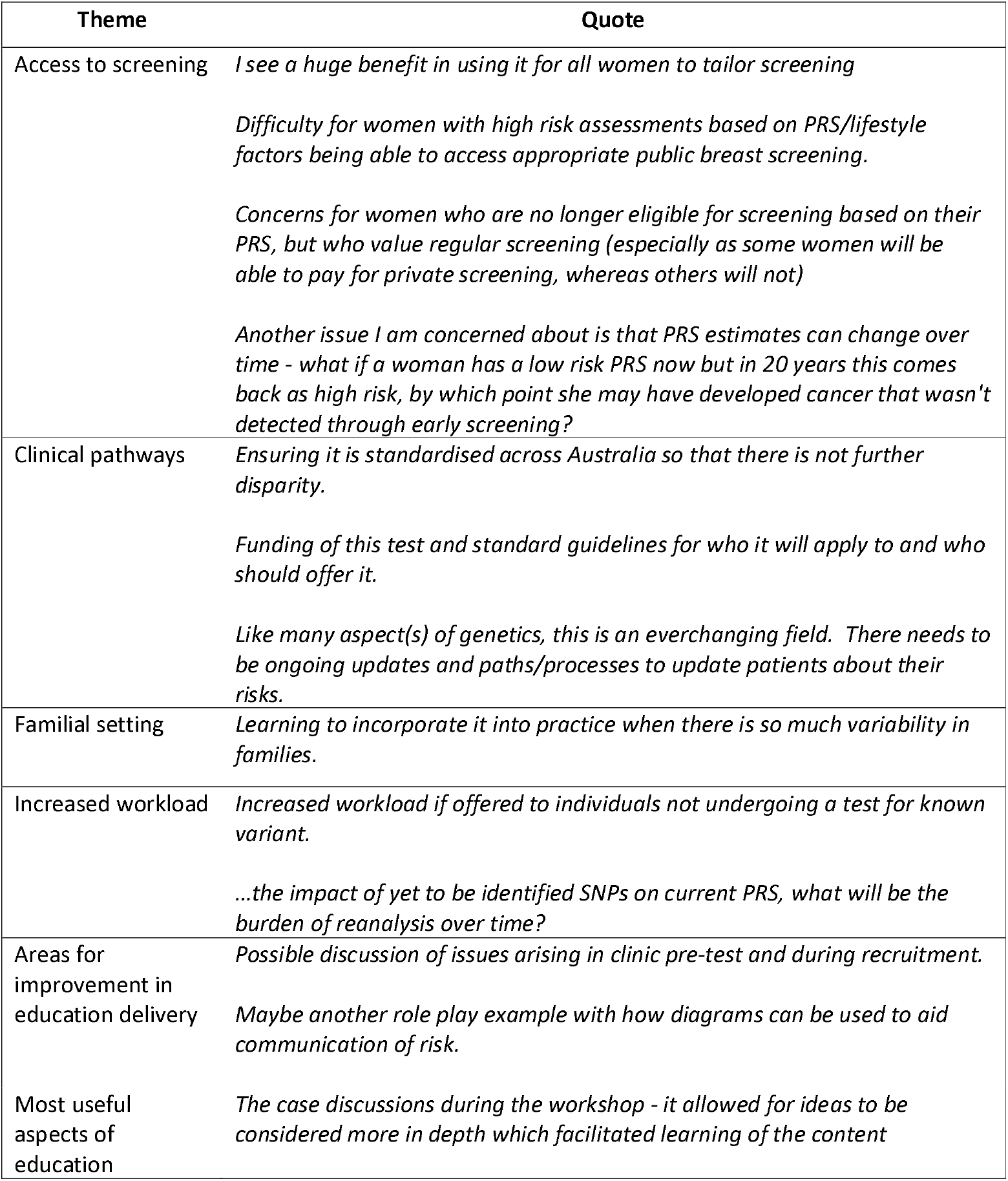
Participants responses to open-ended questions regarding additional challenges to PRS delivery and feedback regarding PRiMo training.

| Theme | Quote |
| --- | --- |
| Access to screening | *I see a huge benefit in using it for all women to tailor screening*<br>*Difficulty for women with high risk assessments based on PRS/lifestyle factors being able to access appropriate public breast screening.*<br>*Concerns for women who are no longer eligible for screening based on their PRS, but who value regular screening (especially as some women will be able to pay for private screening, whereas others will not)*<br>*Another issue I am concerned about is that PRS estimates can change over time - what if a woman has a low risk PRS now but in 20 years this comes back as high risk, by which point she may have developed cancer that wasn't detected through early screening?* |
| Clinical pathways | *Ensuring it is standardised across Australia so that there is not further disparity.*<br>*Funding of this test and standard guidelines for who it will apply to and who should offer it.*<br>*Like many aspect(s) of genetics, this is an everchanging field. There needs to be ongoing updates and paths/processes to update patients about their risks.* |
| Familial setting | *Learning to incorporate it into practice when there is so much variability in families.* |
| Increased workload | *Increased workload if offered to individuals not undergoing a test for known variant.*<br>*...the impact of yet to be identified SNPs on current PRS, what will be the burden of reanalysis over time?* |
| Areas for improvement in education delivery | *Possible discussion of issues arising in clinic pre-test and during recruitment.*<br>*Maybe another role play example with how diagrams can be used to aid communication of risk.* |
| Most useful aspects of education | *The case discussions during the workshop - it allowed for ideas to be considered more in depth which facilitated learning of the content* |

### Confidence and preparedness

Pre-education, participants frequently self-rated as not at all confident/not confident across all eight items measured, most commonly “*interpreting a PRS report*“ (n = 45, 56%), “*discussing insurance implications of PGS*“ (n= 43, 54%), and “*responding to patients’ questions about PGS*“ (n=40, 50%) (Figure 3). Compared to pre-education (M 14.0, SD 6.8), participants were significantly more likely have greater confidence using PRS after completing the educational program (M=23.6, SD 4.7) (t (51)= 13.1, p=<0.001). Areas where lack of confidence remained for some providers post-education were: “*discussing insurance implications of PRS*“, “*making risk management recommendations based on PRS*“, “*responding to a patient’s questions about PRS*“ and “*interpreting a PRS report*“, with 21%, 5%, 3,%, 2% and 2% of participants indicating not at all confident/not confident, respectively. For the remaining four items, no participants indicated not being confident performing those tasks post-training.

**Figure 3:**
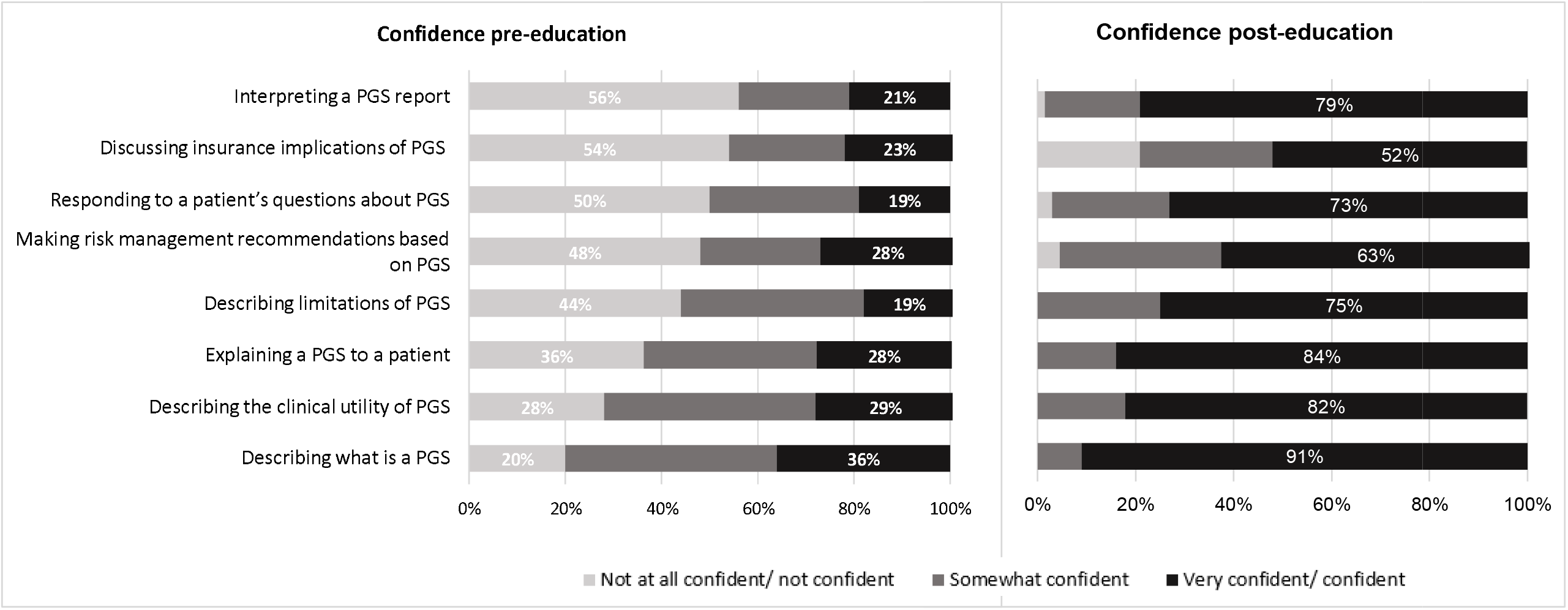
Participants self-reported confidence performing tasks related to PRS pre and post education (n=67)

**Figure 4:**
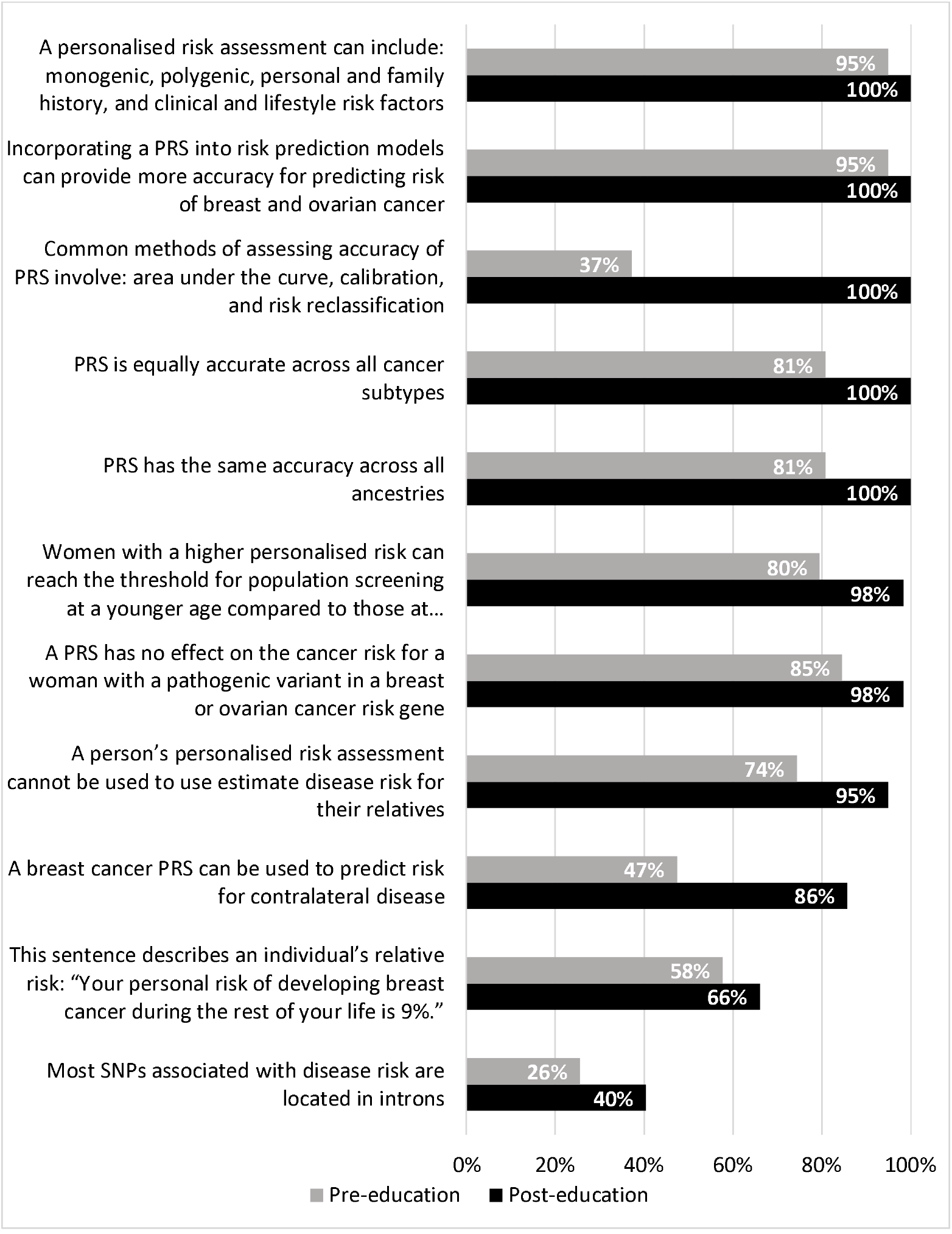
Frequency of correct responses evaluating objective knowledge related to PRS and personalised breast and ovarian cancer risk pre- and (n=80) post-education (n=67)

When asked at baseline how prepared they felt for the integration of PRS and personalized risk in their clinical practice pre-education, about a third of participants indicated feeling not at all prepared (n=30, 38%), followed by 46 (58%) somewhat prepared, and only four (5%) indicated feeling very prepared. Post-education, no participant indicated not being prepared, and the majority indicated being somewhat (n=29, 42%) or very prepared (n=39, 58%). Completion of the education program was associated with significant improvements in preparedness (p=<0.001).

### Knowledge

Pre-education the mean number of correct responses was 7.3 (SD 2.2, range: 1-11). Post-education providers were significantly more likely to correctly answer the knowledge questions (M 9.2, SD 1.3, range 6-11) (t(47)= 7.1, p=>0.001). Questions most frequently answered incorrectly post-education were identifying that most SNPs associated with disease risk are located in intronic regions (n=31, 60%), describing the difference between relative and absolute risk (n=19, 44%), the potential impact of PRS on optimal age for risk-reducing salpingo-oophorectomy for women with a *BRCA1* PV/LPV (n=17, 30%).

### Feedback on educational program

Sixty-seven participants provided feedback on the education program. Most participants indicated that the education program entirely (n=49, 73%) or partially met their learning needs (n=17, 25%). Similarly, most felt the content of the education program was entirely (n=59, 88%) or partially relevant to their clinical practice (n=7, 10%), and that the amount of information provided was about right (n=60, 90%). When asked to rate the most useful aspect of the education program, participants most frequently rated as very/extremely useful the case discussions (n=58, 87%), role plays (n=55, 82%), and educational videos on the website (52, 78%) (Figure 5). This finding is reflected in the responses to the open ended question requesting additional feedback, where 45 providers indicated that they found the role plays and case discussions most valuable as it provided them with an opportunity to reflect on the various issues and discuss the cases with peers and experts. Providers also indicated a desire for ongoing access to the online resource and suggested literature post-training. In relation to areas of improvements, some participants felt that the first two videos of the website were repetitive, and several individuals reported not having had enough time to review the recommended reading prior to the workshop (Table 2). Lastly, the educational program focused entirely on post-test communication. As such, some participants identified a need for further education related to the pre-test consent discussion.

**Figure 5:**
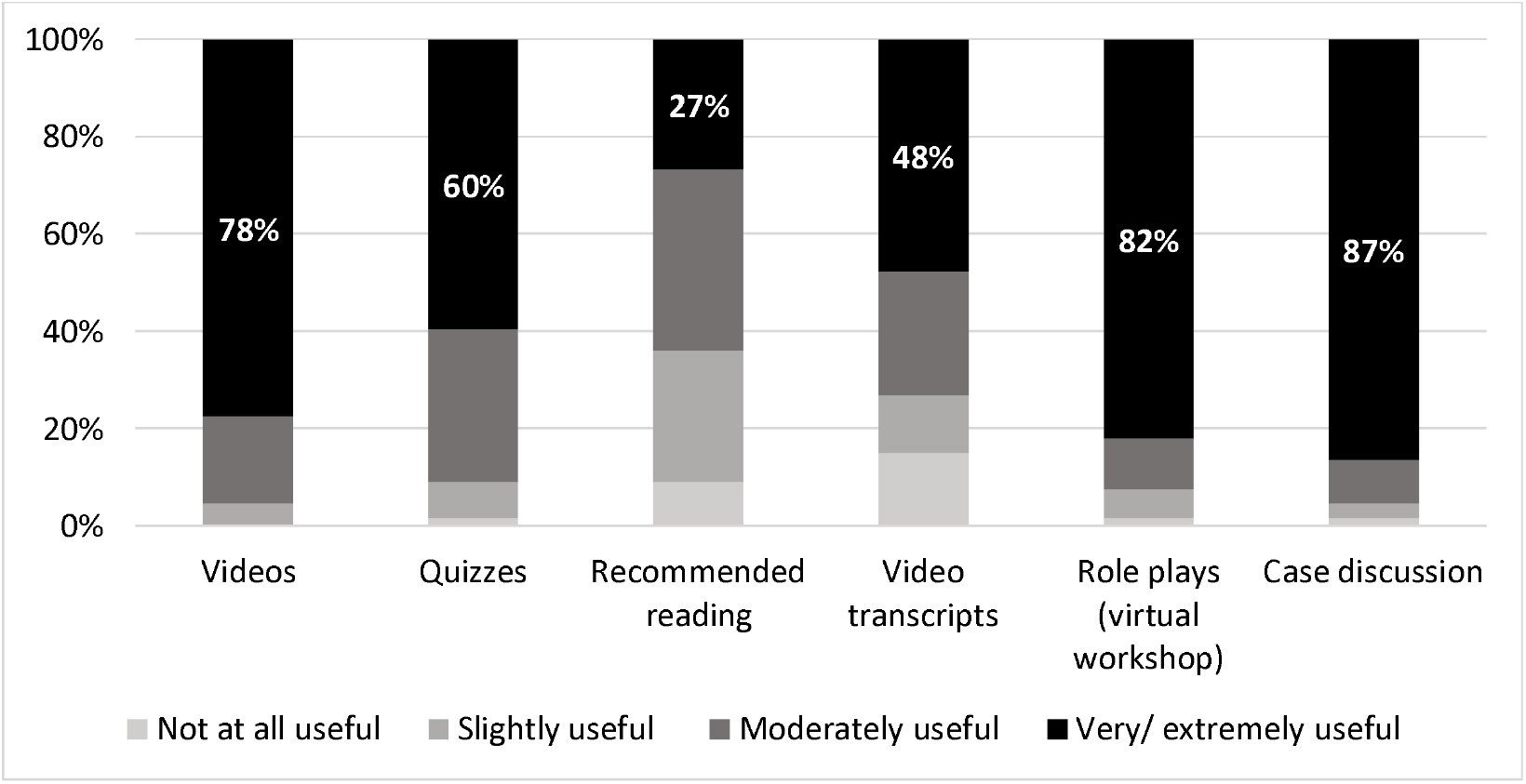
Participants’ perceived usefulness of the education program components (n=67)

## Discussion

Testing for PRS is increasingly being offered through commercial genetic testing companies and clinical research. Yet, to our knowledge, there is no training available for healthcare providers on the use of PRS, representing a significant barrier to implementation of this technology (21-23). To address this issue, we developed and evaluated a novel online education for program for breast and ovarian cancer PRS with 124 genetic healthcare providers across Australia. Most participants had positive reactions to the program, including that their learning needs were met, that the content was relevant to their clinical practice and the amount of information provided was about right. Completion of the training program was also associated with improved attitudes towards, and greater confidence, preparedness, and knowledge of PRS and personalized risk. However, knowledge gaps remained post-training suggesting genetic healthcare providers may require additional support applying PRS in practice.

Delivery of the education program did not alter providers thoughts regarding the potential benefits of providing PRS and personalized risk. This finding is likely due to providers having high endorsement for the benefits of PRS pre-education, including strongly agreeing that PRS can provide greater certainty about risk level and inform risk-management decisions. However, only 64% of the cohort believed that PRS would provide opportunities for reduced screening. Prior studies have identified patient and provider acceptance of reduced screening among those identified to be low risk as a potential barrier to implementation of PRS and risk-stratified screening (31-33). This issue may be exacerbated in the context of monogenic testing and familial risk, particularly in instances where PRS alters risk for family members with the same PV/LPV (16). Currently, familial cancer services primarily focus on monogenic aspects of disease risk, whereas implementation of PRS will require a shift towards personalized model of practice that involves personal evaluation of risk for each individual family member (33, 34). New genetic counselling frameworks and risk communication tools will need to be developed to facilitate discussion of personalized risk information and support patient understanding and acceptance.

In Australia, provision of genetic testing is primarily funded through public healthcare, and eligibility for publicly-funded genetic testing for breast and ovarian cancer is based on established national guidelines (30). Unsurprisingly, our study identified that pre-education most clinicians were already using personalized risk tools in their routine practice to assess eligibility for publicly-funded testing and inform risk management. This finding is of relevance given that implementation of PRS for breast and ovarian cancer risk will occur as part of personalized risk prediction tools, rather than utilizing PRS alone (35). As such, an opportunity exists to demystify and normalize the use PRS by supporting providers to identify areas of practice that they are already familiar with, such as communicating complex risk, evaluating multifactorial disease risk, and using risk prediction tools.

Providers in this study most valued the experiential components of the training i.e., role plays and case discussions with peers and field experts in supporting their learning of PRS and personalized risk. Interactive methods, such as case-based learning, are consistently rated as most preferable by adult learners, and most effective at improving learning, performance, and patient health outcomes (36-38). Benefits of case-based learning include provision of tailored education that is immediately relevant to patients, opportunity for interactions with peers and contextualization of knowledge (36-38). Such benefits are aligned with adult learning theory that posits learners are self-motivated due to a recognized need to learn educational content of perceived relevance to workplace practice (39). Despite the benefits of interactive learning, it is important to consider that this approach is resource intensive, requires administrative support to arrange learning events, expert time to develop case studies and role plays, and moderator support. As PRS availability increases, strategies are needed to upscale PRS training to reach a broader audience, while capturing the benefits of case-based learning and interactive models. One potential approach is developing a “*train-the-trainer*“ model, thereby creating PRS champions who can foster peer learning (38, 40).

Development of this educational program was informed by the Kolb experiential learning model and focused on developing concrete learning, reflective observation, and abstract conceptualization (26). The last item of this model, active experimentation, was not included this educational program. Active experimentation occurs when learners have an opportunity to try what they have learnt, such as seeing patients in a real world setting and discussing PRS information. Active experimentation is an important component of the learning process that involves a gradual building of experience and procedural skills over time (41). Future educational program should consider strategies to support active experimentation, such as online chat groups to provide opportunities to ask questions, identifying PRS champions who can support less experienced staff, and follow-up workshops for reflection and discussion. Additionally, our evaluation only addressed the reaction and learning levels of the Kirkpatrick model. Furthermore, the extent to which the educational program changes practice and behavior beyond the PRiMo study is not known. It is likely that additional implementation barriers, including those identified by study participants (i.e. funding models, lack of clinical practice guidelines and re-analysis over time), will need to be addressed before widespread uptake of PRS in clinical practice. These findings highlight a need for consideration of behavioral change theory to support provider uptake of PRS testing. Theoretical models such as **C**apability, **O**pportunity, **M**otivational model of **B**ehavior change (COM-B) can be used to be used to identify additional barriers to provider uptake of PRS testing and inform development of targeted interventions (42).

Our educational program was targeted to genetic healthcare providers who were participating in a PRS clinical trial, and therefore, had relevant baseline genetic knowledge and were motivated to complete the training. Thus “*just in time*“ training was applied, which is shown to be an efficient method of education (43). To ensure continued learning and successful integration of PRS and personalized risk, a comprehensive suite of educational resources will need to be developed and adapted to different contexts and settings (23). For example, primary care providers and other non-genetic specialists will have different values, learning needs, and perceptions of PRS that will need to be considered in the development of future educational resources. Future needs assessments will need to be conducted to ensure targeted education is developed. Strategies such as point-of-care resources may need to be developed to facilitate patient communication and provider education. Similarly, access to training through other online portals, conference or professional organizations should be considered.

## Conclusions

To date little attention has been paid to healthcare providers’ learning needs regarding PRS and personalized risk. Healthcare providers knowledge and confidence using PRS will inform the successful evaluation and implementation of this new technology. We identified that genetic healthcare providers had little experience using PRS and personalized risk. However, providers frequently described possible benefits of providing this information to patients, including the potential for improved access to tailored screening. Our study demonstrated that implementation of a virtual educational program can significantly improve providers general attitudes, confidence, knowledge, and preparedness for using PRS and personalized risk for breast and ovarian cancer risk. As such we provided a framework for the development of further educational programs to support and upskill providers and thus, facilitate the continued clinical evaluation of PRS and translation to clinical practice.

## Supporting information

Manuscript Supplementary Material

## Data Availability

The anonymized survey data collected and analysed in this study are available from the corresponding author on reasonable request.

## List of abbreviations

PV/LPV: pathogenic/likely pathogenic variants
SNPs: single nucleotide polymorphisms
PRS: polygenic risk score
PRiMo: Using Polygenic Risk Modification to Improve Breast Cancer Prevention

## Declarations

### Ethics approval and consent to participate

This study was approved by the University of Queensland Human Research Ethics Committee (2021/HE001244). Participation in this study was voluntary and informed consent was implied by completion of the anonymous online survey.

### Consent for publication

#### Availability of data and materials

The anonymized survey data collected and analyzed in this study are available from the corresponding author on reasonable request.

### Authors’ contributions

Development of the education program was led by author TY, PAJ, and in consultation with a multidisciplinary team of healthcare professionals and science educators (authors: CW, AML, AW, LM, BT, SM, RW, HK, and MAY). TY led data collection and analysis, with support from CW. SM led participant recruitment and enrollment in the PRS training. All authors contributed to the study design and to the writing of the manuscript. All authors read and approved the final manuscript.

## Acknowledgements

We wish to thank all the familial cancer clinics and genetic healthcare providers who completed the educational program and participated in the study.

