## Supplementary material for "Development and evaluation of a novel educational program for providers on the use of polygenic risk scores": Manuscript Supplementary Material

### Supplementary 1: PRiMo Sites

**1. Department of Medical Genomics**

Royal Prince Alfred Hospital

Camperdown NSW 2050

**2. Familial Cancer Service**

Royal North Shore Hospital

St Leonards NSW 2065

**3. Familial Cancer Service**

Westmead Hospital

Westmead NSW 2145

**4. Hereditary Cancer Centre**

Prince of Wales Hospital

Randwick NSW 2031

**5. Genetic Health Queensland**

Royal Brisbane & Women’s Hospital

Herston QLD 4029

**6. Adult Genetics Unit**

Royal Adelaide Hospital

Adelaide SA 5000

**7. Tasmanian Clinical Genetics Service**

Royal Hobart Hospital

Hobart TAS 7000

**8. Clinical Genetics Service**

Austin Health

Heidelberg VIC 3084

**9. Family Cancer Clinic**

Cabrini Health

Malvern VIC 3144

**10. Familial Cancer Centre**

Monash Health

Clayton VIC 3168

**11. Parkville Familial Cancer Centre**

*Including Barwon Health Familial Cancer Clinic*

Peter MacCallum Cancer Centre & Royal Melbourne Hospital

Melbourne VIC 3000

**12. Familial Cancer Program**

Genetic Services of Western Australia

King Edward Memorial Hospital

Subiaco WA 6008

### Supplementary 2: Study and Education Schematic

1

**Identification:**

Clinician identified by local site lead investigator and contact information sent to the PRiMo study co-ordinator (author SM)

**Recruitment and pre-education data collection:**

Clinician sent email by study co-ordinator (author SM) with:

- link to education website
- invitation to complete anonymous pre-training survey.

**Education stage 1:**

Clinician given two weeks to review online content covering:

i. PRS for Breast and Ovarian Cancer Risk
ii. Personalised Breast and Ovarian Cancer Risk
iii. Breast and Ovarian Cancer Risk Factors
iv. the PRiMo Trial
v. Communicating Personalised Risk Information.

**Education stage 2:**

Clinician attends online virtual workshop moderated by authors TY and PAJ

i. strategies for communicating complex risk information (i.e., monogenic, polygenic and other risk factors)
ii. impact of personalised risk on cancer risk management
iii. communicating personalised risk in the context of familial testing
iv. addressing diversity issues in genomics, PRS and healthcare.

**Post-education data collection:**

Link to post-education survey provided at the end of workshop and survey reminder sent by study co-ordinator one-week post-education. All clinicians were eligible to complete the post-education survey, regardless of whether they completed the pre-survey.

### Supplementary Material 3: Study Measures

| **Measure** | **Description** | **Pre-education** | **Post-education** |
| --- | --- | --- | --- |
| *Demographic characteristics* | To minimise potential for participant identification among the small cohort of genetic healthcare providers in Australia, only limited demographic information was collected, namely: age range, gender, clinical position, and years of clinical practice. | **X** | **X**  (for participants who did not complete survey 1) |
| *Experience using PRS and personalised risk* | 3-items assessing i) how much information participants previously heard about PRS, ii) whether they ever provided PRS to a patient (either clinically or in research), and iii) prior use of the CanRisk tool. | **X** |  |
| *General attitudes* | 6-items were adapted from McGuinness, Fassi, Wang, Hacking, Ellis ^15^ to assess participant’s attitudes towards PRS, with items measured on a 5-point scale ranging from “*strongly disagree*” to “*strongly agree*”. | **X** | **X** |
| *Perceived benefits and concerns* | 11-items were developed to evaluate level of agreement regarding potential benefits (5 items), and concern for potential negative outcomes (6 items). | **X** | **X** |
| *Confidence* | 8-items were developed to evaluate confidence completing different aspects of using PRS and personalised risk assessments. The confidence items were developed to align with the training learning outcomes and items were scored on a 5-point scale from “*not at all confident*” to “*very confident*”. | **X** | **X** |
| *Preparedness* | 1-item, participants were asked to rate how prepared they were for the integration of this information in clinical practice on a five-point scale ranging from “*not at all prepared*”, to “*very prepared*”. | **X** | **X** |
| *Knowledge of PRS and personalised cancer risk* | 11 true/false questions were developed to assess objective knowledge of PRS and personalised risk. Knowledge questions were developed to align with information provided during the website modules | **X** | **X** |
| *Training evaluation* | 6-items were adapted from the Royal Australian College of General Practitioners guidelines^19^ to evaluate training programs. Questions include extent to which learning needs were met, relevance of education to individual practice and length of website and virtual workshop. Participants were also asked to rate the usefulness of the different aspects of the education program (i.e. videos, quizzes and roleplays). |  | **X** |
| *Open ended questions* | open-ended questions asked participants if they had any additional thoughts regarding the benefits, limitation, and potential concerns related to PGS and personalised risk. | **X** | **X** |
